# Trends and determinants of unmet need for modern contraception among adolescent girls and young women in Tanzania, 2004-2016

**DOI:** 10.1101/2022.06.07.22276109

**Authors:** Sophia Adam Kagoye, Ola Jahanpour, Joseph Obure, Michael Johnson Mahande, Jenny Renju

**Author notes:** **Corresponding author Email:** (SK). These authors contributed equally to this work.

## Abstract

**Background:** Unintended pregnancy at a young age can lead to poor reproductive health, social and economic outcomes. The high rate of unintended teenage pregnancies in Tanzania is indicative of inadequate availability and uptake of modern contraception. Determining trends and determinants of unmet need for modern contraception among adolescent girls and young women (AGYW) in Tanzania will help address the burden of unintended pregnancies.

**Methodology:** An analytical cross-sectional study design was conducted using secondary data from three consecutive Tanzania Demographic and Health Surveys (TDHS) 2004/05, 2010 and 2015/6. Data analysis was performed using Stata version 15.0. Data analysis considered the complex survey design. Categorical and continuous variables were summarized using descriptive statistics. Poisson regression model was used to determine factors associated with unmet need for modern contraception.

**Results:** A steady decline in unmet need for modern contraception was observed from 31.8% in 2004/05 to 27.5% in 2015/16 survey. In the multivariable analysis, higher prevalence of unmet need for modern contraception was observed among adolescents, participants with at least one live birth, from poor wealth tertile, and those sexually active during the past four weeks.

**Conclusion:** Despite declining levels, the unmet need for modern contraception among young women in Tanzania remains high. AGYW under 19 years, those from poor households, and those who are postpartum are most at risk. Greater efforts are required to meet the reproductive health needs and rights of these sub-groups of AGYW in order to facilitate uptake of modern contraceptives and therefore reducing the risk of unintended pregnancies and unmet need for modern contraception.

## Introduction

An unmet need for modern contraception is defined as the proportion of fecund (fertile) sexually active women who want to limit or delay childbearing beyond two years, but are not using modern contraceptive methods (1,2). According to 2016 estimates, only 43% (16 million) of the 36 million sexually active adolescent girls in developing countries were using modern contraceptives leaving the remaining 57% (20 million) with an unmet need (3,4). Nearly half (49%) of the 21 million adolescent (aged 15-19 years) pregnancies in developing countries are unintended and more than half (55%) of unintended pregnancies in developing countries end in unsafe abortions (3).

Early childbearing is associated with an increased risk of maternal mortality and morbidities, such as unsafe abortions, endometriosis, severe pre-eclampsia and preterm labor (3,5–8). It is estimated that in Tanzania between 2004 and 2016, maternal comorbidities accounted for 18.5% of deaths amongst female adolescents (15-19 years) and 31.5% amongst young women (20-24 years) (9). Childbearing at a young age is also linked to higher rates of infant and child mortality (3,5–8). Furthermore, unintended pregnancies among adolescent girls and young women are attributed to increased school dropout and risk of intimate partner violence (physical, sexual, emotional violence) (8,10). Such consequences place a socio and economic burden on individuals, families, communities and the broader health care system (8,10).

The FP2020 Initiative target aims to enable 120 million additional users to take up modern contraceptives in low-income countries by 2020(11,12). In order for Tanzania to reach their targets under this initiative, national policy and guidelines have been developed to promote the implementation of acceptable, accessible, comprehensive, effective, efficient, equitable and appropriate adolescent and youth friendly sexual and reproductive health services. These services provide an opportunity for adolescents and youths to access free contraceptive services, gain family planning (FP) education and comprehensive sexual education in health facilities from a primary level to tertiary level (13–17).

Despite these efforts, the proportion of AGYW with an unmet need for modern contraceptives remains unacceptably high and persistent (ranging from 15.6% in 2004/05 to 14% in 2010 and 2015/16 respectively)(9,18,19). Consequently, the rates of unintended pregnancies in Tanzania among adolescent girls have increased from 10.4% in 2004/5 to 32.6% in 2015/6 and from 21.8% to 30.7% for young women in the same period (9,18,19). Consequently, adolescent fertility rate targets of less than 100 per 1000 live births by 2020 have not been met.

Little is known as to what factors influenced the observed changes in the unmet need for modern contraception among AGYW and how these associated factors might have changed over time in this population. Understanding these factors will support the development of evidence-based targeted interventions designed to meet the contraceptive needs of AGYW to ultimately reduce unintended pregnancies. This study aimed to determine trends and determinants of changes in unmet need for modern contraception among adolescent girls and young women in Tanzania.

## Materials and methods

### Study design and setting

We conducted an analytical cross-sectional study using nationally representative data from three consecutive Tanzania demographic and health surveys (TDHS) (2004/05, 2010 and 2015/16). The TDHS includes data from all regions of the United Republic of Tanzania and employs a multi-stage sampling procedure, the TDHS and its methods have been described in detail elsewhere (20). In brief, stratification into homogenous subgroups/strata from the sampling frame typically geographical areas and urban/rural residence was conducted in order to minimize sampling error. The first sampling stage involves a selection of primary sampling units (PSUs) with probability proportion to size within each stratum, these are typically census enumeration areas (EAs) and form the survey clusters. In the second stage, using a complete household listing in each of the selected clusters, a fixed number of households is selected by equal probability systematic sampling techniques. The overall selection probability for each household is the probability of selecting the cluster, multiplied by the probability of selecting the household within the cluster(20).

### Study population and sample size

This study included all married and unmarried AGYW aged 15-24 years who reported to have ever had sex. Individuals with missing information on sexual activity, fertility desires on wantedness of births or a current pregnancy were excluded. Also, individuals with a desire for a child within 24 months and those who were infecund were excluded. AGYW were classified as infecund if they fell into any of the following categories: (a) married 5+ years, had no children in the past 5 years and never used contraception; (b) responded “can’t get pregnant” to question on desire for future children; (c) responded “hysterectomy” on reason for not using contraception; (d) response to time since last period is > 6 months and did not report postpartum amenorrhea and (e) response to time since last period is “never menstruated” or “hysterectomy”.

The total weighted sample size from the 3 surveys was 7,525 AGYW (aged 15 – 24years) whereby 2,286 adolescent girls and young women were from the 2004-2005 survey, 2,218 adolescent girls and young women from the 2010 survey and 3,021 adolescent girls and young women from the 2015-2016 survey respectively (Fig **1**).

**Fig 1.**
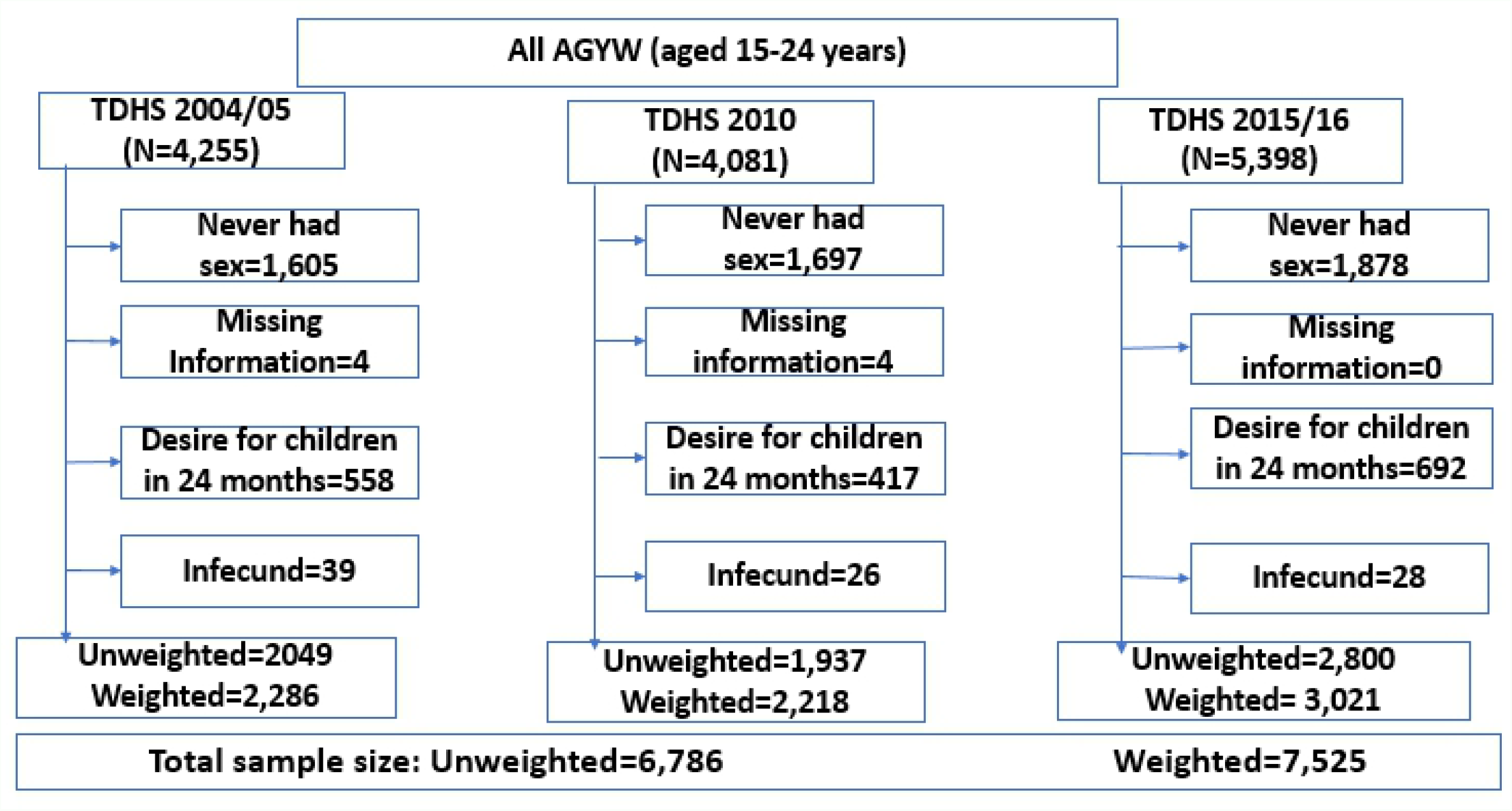
Flow chart for selection of study participants.

### Measures

#### Dependent variable

The dependent variable in this study was an **unmet need for modern contraception** which was categorized dichotomously as “Yes/No”. Modern contraceptive methods included the pill, injectables, intrauterine devices, implants, female and male sterilization, female and male condoms, other barrier methods, and modern fertility-awareness methods(21). In this study we adopted a revised DHS definition(22) to calculate the dependent variable. Specifically, married or ever had sex yet unmarried AGYW were considered to have unmet need for modern contraception if they were: (a) fecund and want a child in 2+ years or want another child but are undecided on the timing or are undecided if they want another child; (b) fecund and do not want any more children; (c) currently pregnant but wanted the current pregnancy later or did not want it at all; (d) postpartum amenorrhoeic and wanted the last birth later or did not want it at all. (AGYW were considered to be postpartum amenorrhoeic if their period had not returned since their last live birth in the past 24 months prior to the survey).

#### Independent variables

The independent variables were grouped into socio-demographic characteristics and reproductive health characteristics.

Specifically, socio-demographic characteristics included: age in years (15-19, 20-24), area of residence (urban or rural), geographical zone (Western, Northern, Central, Southern Highlands, Southern, South West Highlands, Lake, Eastern, Zanzibar), marital status (unmarried, married/cohabiting), woman’s education level (no education, primary education, secondary & above), currently working (yes or no), wealth index tertiles (poor, middle, rich) and family planning media exposure (yes or no).

Reproductive health characteristics included: age at sexual debut (<15 years, ≥15 years), parity (nulliparous, primiparous or multipara); fertility preference (wanting a child after 2+ years, wanting a child but unsure on the timing, undecided if they want a child, do not want any more children), ever terminated pregnancy (yes or no), sexually active in the past 4 weeks (yes or no), ever in contact with FP field worker (yes or no).

### Statistical Analysis

Data was analyzed using Stata version 15.0 (StataCorp. 2017. Stata Statistical Software: Release 15. College Station, TX: StataCorp LLC). Some variables were categorized based on previous literature for better interpretation and comparability between studies. Descriptive statistics were summarized using frequency and proportions for categorical variables while continuous variables were summarized using measures of central tendency with respective measures of dispersion. Trends in unmet need for modern contraception and modern contraceptive use were computed as proportions. Differences in unmet need by background and reproductive health characteristics were further examined and percentage point changes were computed. The trends were examined separately in three phases 2004/05-2010 as Phase1 (survey 1 to survey 2), 2010-2015/16 as Phase II (survey 2 to survey 3) and 2004/05-2015/16 as Phase III (survey 1 to survey 3).

Classical multivariable logistic regression was initially considered to determine factors associated with unmet need for modern contraception. However, this approach was discarded due the drawbacks of logistic regression of over-estimating odds ratios and 95% confidence intervals for outcomes with a prevalence greater than 10%. Since in our case the prevalence of unmet need for modern contraception was over 10% for all the 3 surveys, classical logistic regression was not used and other alternatives to classical logistic regression were considered.

A Log-binomial regression was considered to determine the factors associated with unmet need for modern contraception. However, this approach also was discarded due to its non-convergence problem (failed to converge). Modified Poisson regression analysis could not be performed since the data were already set to account for complex survey design and the option *vce (robust)* was not available. Therefore, Poisson regression model was used to determine the factors associated with unmet need for modern contraception.

Crude and adjusted analysis were performed separately for each survey year to determine the changes in prevalence ratios of the exposures on unmet need for modern contraception. The magnitude of association was interpreted using the prevalence ratios with their corresponding 95% Confidence Interval, A *p*-value of less than 0.05 (2-tails) was considered statistically significance.

All estimates took into account the complex survey nature of the data thus weighting for sampling probabilities and non-response, clustering and stratification done in the TDHS, (*svy*) STATA command was used accordingly.

### Ethical Consideration

Ethical approval to carry out the study was obtained from Tumaini University, KCMUCo Research and Ethical Committee (# PG016/2019). Permission to use DHS data was granted from the DHS MEASURE program. The TDHS has ethical approval from ICF Institutional Review Board (IRB) as well as from local IRB in Tanzania and written informed consent was obtained from all the study participants above 18 years. For each participant under 18 years of age, written informed consent was obtained from the parent/guardian. Detailed information on ethical consideration of DHS program are published in DHS reports(9,18,19).

## Results

### Characteristics of the study participants

#### Socio-demographic characteristics of study participants

A total of 7,525 participants across the three surveys were studied, 2004/05 survey (n=2,286), 2010 survey (n=2,218) and 2015/16 survey (n=3,021). The sociodemographic characteristics of the study participants are shown in Table **1**. The median age of study participants across the surveys was 20 (IQR= 18-22), 21 (IQR= 18-23) and 20 (IQR=18-22) years for 2004/05, 2010 and 2015/16 surveys respectively. Across the three survey rounds the majority of the study participants were aged between 20 to 24 years, and their proportion slightly decreased over time from 63.6% in 2004/05 to 62.9% in 2010 and 59.6% in 2015/16. The majority were from rural areas (70%, 68.6% and 62.8% for 2004/05, 2010 and 2015/16 surveys respectively). More than half (63.7% in 2004/05 to 55.2% in 2010 then to 52.6% in 2015/16) of the study participants were married. The proportion of adolescent girls and young women with secondary education and above increased over time from 8.5% in 2004/05 to 21.1% 2010 and 31.3% in 2015/16 surveys respectively. The proportion of adolescent girls and young women with FP media exposure declined from 61.6% in 2004/05 to 53.8% in 2010 and then increased to 69.5% in 2015/16 **(Table 1)**.

**Table 1:** Socio-demographic characteristics of study participants in TDHS 2004/2005, 2010 and 2015/2016 surveys (N=7,525)

| Variable | Survey Year |  |  |
| --- | --- | --- | --- |
|  | TDHS 2004/05<br>(n= 2,286)<br>n (%) | TDHS 2010<br>(n=2,218)<br>n (%) | TDHS 2015/16<br>(n=3,021)<br>n (%) |
| **Age** |  |  |  |
| 15-19 | 831 (36.4) | 822 (37.1) | 1,222 (40.4) |
| 20-24 | 1,455 (63.6) | 1,396 (62.9) | 1,799 (59.6) |
| *Median (IQR)* | *20 (18,22)* | *21 (18, 23)* | *20 (18,22)* |
| **Area of residence** |  |  |  |
| Urban | 687 (30.0) | 697 (31.4) | 1,125 (37.2) |
| Rural | 1,599 (70.0) | 1,521 (68.6) | 1,896 (62.8) |
| **Zone** |  |  |  |
| Western | 210 (9.2) | 221 (10.0) | 285 (9.4) |
| Northern | 239 (10.5) | 240 (10.8) | 298 (9.9) |
| Central | 219 (9.6) | 197 (8.9) | 295 (9.8) |
| Southern Highlands | 179 (7.8) | 175 (7.9) | 173 (5.7) |
| Southern | 142 (6.2) | 117 (5.3) | 170 (5.6) |
| South West Highlands | 216 (9.4) | 176 (7.9) | 303 (10.0) |
| Lake | 656 (28.7) | 656 (29.6) | 899 (29.8) |
| Eastern | 390 (17.1) | 404 (18.2) | 555 (18.4) |
| Zanzibar | 35 (1.5) | 32 (1.4) | 43 (1.4) |
| **Marital Status** |  |  |  |
| Unmarried | 830 (36.3) | 993 (44.8) | 1,432 (47.4) |
| Married/Cohabiting | 1456 (63.7) | 1,225 (55.2) | 1,589 (52.6) |
| **Education level** |  |  |  |
| No education | 528 (23.1) | 398 (17.9) | 254 (8.4) |
| Primary education | 1,563 (68.4) | 1,353 (61.0) | 1,821 (60.3) |
| Secondary and above | 195 (8.5) | 467 (21.1) | 945 (31.3) |
| **Currently working** |  |  |  |
| No | 564 (24.7) | 602 (27.3) | 1,110 (36.4) |
| Yes | 1,719 (75.3) | 1,604 (72.7) | 1,919 (63.6) |
| **Wealth index** |  |  |  |
| Poor | 835 (36.5) | 767 (34.6) | 1,067 (35.3) |
| Middle | 425 (18.6) | 412 (18.6) | 519 (17.2) |
| Rich | 1,026 (44.9) | 1,039 (46.8) | 1,434 (47.5) |
| **FP media exposure <sup>a b</sup>** |  |  |  |
| No | 877 (38.4) | 1,023 (46.2) | 922 (30.5) |
| Yes | 1,408 (61.6) | 1,194 (53.8) | 2,099 (69.5) |

IQR=Interquartile range; <sup>a</sup> 2004/05 n=2,285; <sup>b</sup> 2010 n=2,217

#### Reproductive Health Characteristics of study participants

The mean age at sexual debut across the three surveys was 16 (SD=2) years. Majority reported their age at sexual debut to be greater than 15 years (59.9% in 2004/05 survey, 61.5% in 2010 survey and 62.4% in 2015/16, survey). The majority of adolescent girls and young women across the survey years reported wanting a child after more than two years (74.1, 73.2% and 79.1% in 2004/05, 2010 and 2015/16 surveys respectively). The proportion of adolescent girls and young women who had ever terminated pregnancy or had abortion or stillbirth increased from 6.9% in 2004/05 to 8.6% in 2010 and then declined to 6.3% in 2015/16. The proportion of those that were sexually active during the past 4 weeks prior the survey decreased over time from 54.5% to 51.4% to 48.6% in 2004/05, 2010 and 2015/16 surveys respectively. The majority of participants had a good knowledge of modern contraceptives and their proportion increased over time from 97% in 2004/05 to 98.3% in 2010 and 98.7% in 2015/16 surveys respectively. The proportion of adolescent girls and young women who reported to have been in contact with a family planning worker either in the households or in health facilities during the past 12 months increased from 27.8% in 2004/05 to 30.4% in 2010 and then decreased to 27.1% in 2015/16 surveys respectively (Table **2**).

**Table 2:** Reproductive health Characteristics of study participants in TDHS 2004/2005, 2010 and 2015/2016 surveys (N=7,525)

| Variable | Survey years |  |  |
| --- | --- | --- | --- |
|  | TDHS 2004/05<br>n=2,286<br>n (%) | TDHS 2010<br>n=2,218<br>n (%) | TDHS 2015/16<br>n=3,021<br>n (%) |
| **Age at sexual debut<sup>a b c</sup>** |  |  |  |
| ≤15 years | 905 (40.1) | 839 (38.5) | 1,130 (37.6) |
| >15 years | 1,351 (59.9) | 1,338 (61.5) | 1,878 (62.4) |
| *Mean ±SD* | *16 ±2* | *16 ±2* | *16 ±2* |
| **Parity** |  |  |  |
| Nulliparous | 626 (27.4) | 722 (32.6) | 988 (32.7) |
| Primiparous | 856 (37.4) | 726 (32.7) | 1,225 (40.5) |
| Multipara | 804 (35.2) | 770 (34.7) | 809 (26.8) |
| **Fertility Preference** |  |  |  |
| Want child after 2+ years | 1,712 (74.9) | 1,621 (73.2) | 2,388 (79.1) |
| Wants child, unsure timing | 224 (9.8) | 400 (18.0) | 335 (11.1) |
| Undecided | 121 (5.3) | 42 (1.9) | 158 (5.2) |
| Want no more | 228 (10.0) | 154 (6.9) | 140 (4.6) |
| **Pregnancy terminations**<br>**(abortion, stillbirths)** |  |  |  |
| No | 2,129 (93.1) | 2,027 (91.4) | 2,829 (93.7) |
| Yes | 157 (6.9) | 191 (8.6) | 192 (6.3) |
| **Sexually active past 4 weeks** |  |  |  |
| No | 1,041 (45.5) | 1,079 (48.6) | 1,553 (51.4) |
| Yes | 1,245 (54.5) | 1,139 (51.4) | 1,468 (48.6) |
| **Ever in contact with FP**<br>**worker (past 12 months)** |  |  |  |
| No | 1,651 (72.2) | 1,543 (69.6) | 2,203 (72.9) |
| Yes | 635 (27.8) | 675 (30.4) | 818 (27.1) |

SD- Standard Deviation; <sup>a</sup> 2004/05 n=2,256; <sup>b</sup> 2010 n=2,177; <sup>c</sup> 2015/16 n=3,008

### Trends in the unmet need for modern contraception

#### Overall trends across the survey years

The total unmet need for modern contraception among sexually active AGYW declined over time from 31.8% to 28.6% to 27.5% in 2004/05, 2010 and 2015/16 surveys respectively. The largest decline of 4.3% in the unmet need for modern contraception was observed between 2004/05 to 2015/16 compared to 3.2% decline between 2004/05 to 2010 and 1.1% decline between 2010 to 2015/16 respectively (Fig **2**).

**Fig 2:**
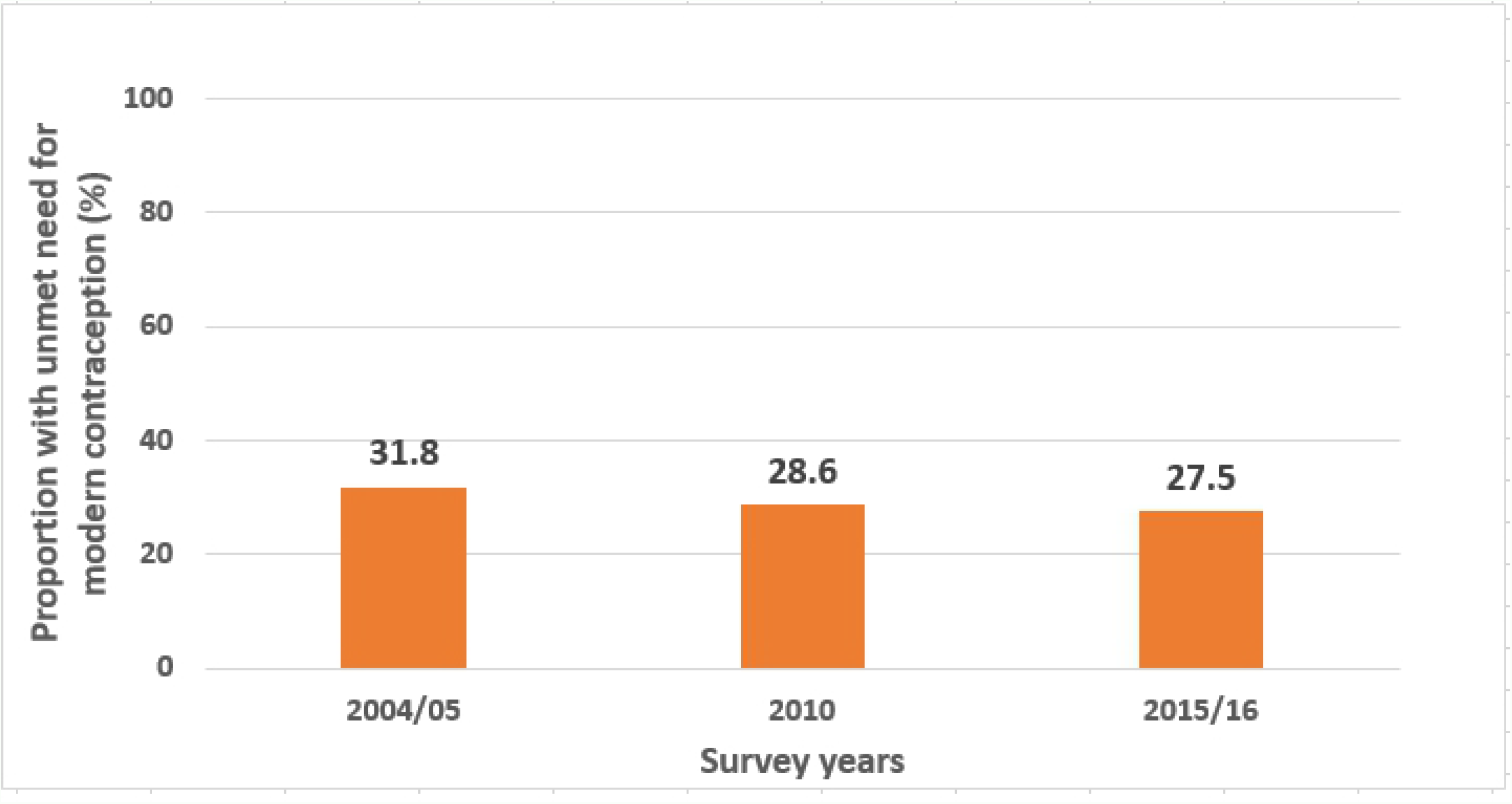
Trends in unmet need for modern contraception among sexually active adolescent girls and young women in Tanzania (TDHS 2004/05-2015/16)

### Trends in unmet need for modern contraception by background characteristics

The proportion of adolescent girls and young women with an unmet need for modern contraception varied by various socio-demographic and reproductive health characteristics as presented in Table **3**.

**Table 3:** Trends in unmet need for modern contraception among sexually active adolescent girls and young women background characteristics, in the TDHS 2004/2005, 2010 and 2015/2016 surveys (N=7,525)

| Characteristic | TDHS<br>2004/2005<br>(n=2,286) | TDHS<br>2010<br>(n=2,218) | TDHS<br>2015/2016<br>(n=3,021) | % point difference in<br>unmet need for<br>modern contraception |  |  |
| --- | --- | --- | --- | --- | --- | --- |
|  |  |  |  | Phase<br>I <sup>a</sup> | Phase<br>II <sup>b</sup> | Phase<br>III <sup>c</sup> |
| **Age** |  |  |  |  |  |  |
| 15-19 | 32.2 | 25.9 | 25.8 | -6.3 | -0.1 | -6.4 |
| 20-24 | 28.3 | 24.9 | 27.0 | -3.4 | 2.1 | -1.3 |
| **Area of residence** |  |  |  |  |  |  |
| Urban | 25.6 | 22.7 | 23.9 | -2.9 | 1.2 | -1.7 |
| Rural | 32.1 | 26.4 | 28.1 | -5.7 | 1.7 | -4.0 |
| **Zone** |  |  |  |  |  |  |
| Western | 33.4 | 26.6 | 25.4 | -6.8 | -1.2 | -8.0 |
| Northern | 14.6 | 24.0 | 23.6 | 9.4 | -0.4 | 9.0 |
| Central | 44.3 | 30.6 | 18.6 | -13.7 | -12.0 | -25.7 |
| Southern Highlands | 22.7 | 21.1 | 27.9 | -1.6 | 6.8 | 5.2 |
| Southern | 35.3 | 31.9 | 15.6 | -3.4 | -16.3 | -19.7 |
| South West Highlands | 13.3 | 30.9 | 28.9 | 17.6 | -2.0 | 15.6 |
| Lake | 35.4 | 26.3 | 34.0 | -9.1 | 7.7 | -1.4 |
| Eastern | 26.3 | 18.6 | 20.8 | -7.7 | 2.2 | -5.5 |
| Zanzibar | 51.2 | 26.6 | 32.4 | -24.6 | 5.8 | -18.8 |
| **Marital Status** |  |  |  |  |  |  |
| Unmarried | 28.8 | 25.2 | 21.8 | -3.6 | -3.4 | -7.0 |
| Married/Cohabiting | 32.4 | 26.0 | 34.9 | -6.4 | 8.9 | 2.5 |
| **Education level** |  |  |  |  |  |  |
| No education | 35.0 | 34.5 | 24.0 | -0.5 | -10.5 | -11.0 |
| Primary education | 29.4 | 27.2 | 29.3 | -2.2 | 2.1 | -0.1 |
| Secondary and above | 18.7 | 17.8 | 20.3 | -0.9 | 2.5 | 1.6 |
| **Currently working** |  |  |  |  |  |  |
| No | 30.9 | 19.9 | 27.1 | -11.0 | 7.2 | -3.8 |
| Yes | 29.3 | 28.2 | 26.1 | -1.1 | -2.1 | -3.2 |
| **Wealth index** |  |  |  |  |  |  |
| Poor | 37.0 | 33.2 | 32.9 | -3.8 | -0.3 | -4.1 |
| Middle | 30.1 | 20.8 | 22.8 | -9.3 | 2.0 | -7.3 |
| Rich | 24.4 | 22.1 | 22.7 | -2.3 | 0.6 | -1.7 |
| **FP media exposure** |  |  |  |  |  |  |
| No | 34.8 | 28.2 | 28.6 | -6.6 | 0.4 | -6.2 |
| Yes | 26.5 | 22.7 | 25.5 | -3.8 | 2.8 | -1.0 |
| **Age at sexual debut** |  |  |  |  |  |  |
| ≤15 years | 33.2 | 27.6 | 29.5 | -5.6 | 1.9 | -3.7 |
| >15 years | 26.2 | 23.3 | 23.4 | -2.9 | 0.1 | -2.8 |

<sup>a</sup> 2004/05-2010; <sup>b</sup> 2010- 2015/16; <sup>c</sup>2004/05-2015/16

**Table 3 (continued): Trends in unmet need for modern contraception among sexually active adolescent girls and young women by background characteristics, in the TDHS 2004/2005, 2010 and 2015/2016 surveys (N=7,525)**
| Characteristic | TDHS<br>2004/05<br>(n=2,286) | TDHS<br>2010<br>(n=2,218) | TDHS<br>2015/2016<br>(n=3,021) | Percentage point<br>difference in unmet<br>need for modern<br>contraception |  |  |
| --- | --- | --- | --- | --- | --- | --- |
|  |  |  |  | Phase<br>I <sup>a</sup> | Phase<br>II <sup>b</sup> | Phase<br>III <sup>c</sup> |
| **Parity** |  |  |  |  |  |  |
| Nulliparous | 23.5 | 18.2 | 17.7 | -5.3 | -0.5 | -5.8 |
| Primiparous | 37.2 | 35.8 | 37.3 | -1.4 | 0.1 | 0.1 |
| Multipara | 42.6 | 39.4 | 39.0 | -3.2 | -3.6 | -3.6 |
| **Fertility preference** |  |  |  |  |  |  |
| Want child after 2+ years | 26.6 | 24.5 | 28.5 | -2.1 | 4.0 | 1.9 |
| Wants child, unsure timing | 25.2 | 25.9 | 15.6 | 0.7 | -10.3 | -9.6 |
| Undecided | 48.2 | 26.6 | 28.8 | -21.6 | 2.2 | -19.4 |
| Want no more | 51.0 | 33.0 | 42.7 | -18.0 | 9.7 | -8.3 |
| **Pregnancy termination**<br>**(abortion, stillbirth)** |  |  |  |  |  |  |
| No | 30.2 | 25.8 | 26.4 | -4.4 | 0.6 | -3.8 |
| Yes | 25.9 | 7.2 | 28.3 | -18.7 | 21.1 | 2.4 |
| **Sexually active past 4**<br>**weeks** |  |  |  |  |  |  |
| No | 23.9 | 20.0 | 20.3 | -3.9 | 0.3 | -3.6 |
| Yes | 38.5 | 36.7 | 35.1 | -1.8 | -1.6 | -3.4 |
| **Ever in contact with a FP**<br>**worker (past 12 months)** |  |  |  |  |  |  |
| No | 29.8 | 23.8 | 24.2 | -6.0 | 0.4 | -5.6 |
| Yes | 30.6 | 31.5 | 35.0 | 0.9 | 3.5 | 4.4 |
| **Total** | **31.8** | **28.6** | **27.5** | **-3.2** | **-1.1** | **-4.3** |

<sup>a</sup> 2004/05-2010; <sup>b</sup> 2010- 2015/16; <sup>c</sup>2004/05-2015/16

Disaggregating the age groups, the total unmet need for modern contraception among adolescent girls (15-19 years) was observed to decline from 30% in 2004/05 to 25.3% in 2010 and then increased to 26.5% in 2015/16 survey while the total unmet need for modern contraceptive among young women (20-24 years) declined across the survey rounds from 32.9% to 30.5% then to 28.1% in 2004/05, 2010 and 2015/16 surveys respectively.

Trends of unmet need for modern contraception also varied by marital status. Unmet need for modern contraception was observed to be predominantly higher among married/cohabiting adolescents compared to unmarried adolescents across the three survey rounds.

There was a decline in unmet need for modern contraception among AGYW by residence (urban/rural) between 2004/05 to 2015/16 with a larger decline of 4% observed among AGYW living in rural areas compared to 1.7% decline observed among those living in urban areas during the same period. We also observed a 25.7% decline in the unmet need for modern contraception from 44.3% 2004/05 to 18.6% in 2015/16 among AGYW living in the Central zone compared to other geographical zones.

### Factors associated with unmet need for modern contraception among sexually active AGYW in Tanzania (2004/05 to 2015/16)

#### Crude analysis

In the crude analysis, across the three survey rounds, we observed a high prevalence of unmet need for modern contraception among adolescent girls and young women living in rural areas, married, from a poor and middle wealth quintile, with no FP media exposure, with age at sexual debut below or equal to 15 years, with at least one live birth (primiparous and multipara), wanting no more children and those that were sexually active during the past 4 weeks prior the surveys.

#### Adjusted analysis

In the adjusted analysis, adolescent girls (15-19 years), adolescent girls and young women from a poor wealth quintile, with at least one live birth (primiparous, multipara) wanting no more children and sexually active during the past 4 weeks prior the surveys were observed to have a significant higher prevalence of unmet need for modern contraception across the three survey rounds.

The effect of age was observed to be significant in 2004/05 and 2015/16 survey rounds. Whereby, after adjusting for other factors adolescent girls (15-19 years) had a 28% and 27% higher prevalence of unmet need for modern contraception compared to young women (20-24) in 2004/05 (APR: 1.28; 95% CI: 1.06, 1.55) and 2015/16 (APR: 1.27; 95% CI: 1.07, 1.51) surveys respectively. During 2010, although not significant, adolescent girls (15-19 years) had 24% higher prevalence of unmet need for modern contraception compared to young women (APR: 1.24; 95% CI: 0.99, 1.55).

The effect of wealth index was observed to increase over time and remained significant in 2010 and 2015/16 surveys. After adjusting for other factors, adolescent girls and young women from a poor wealth quintile had 32% and 46% significantly higher prevalence of unmet need for modern contraception compared to those from a rich quintile in 2010 (APR: 1.32 ; 95% CI: 1.03, 1.69) and 2015/16 (APR: 1.46; 95% CI: 1.18, 1.81) surveys respectively. In 2004/05, adolescent girls and young women had 16% higher prevalence of unmet need for modern contraception compared to those in the rich quintile (APR: 1.16; 95% CI: 0.94, 1.43).

The effect of parity remained significant across the three survey rounds. After adjusting for other factors, adolescent girls and young women with one livebirth (primiparous) had 79%, 84% and 68% significantly higher prevalence of unmet need for modern contraception compared to those with no livebirth (nulliparous) in 2004/05 (APR: 1.79; 95% CI: 1.42, 2.27), 2010 (APR: 1.84; 95% CI: 1.39, 2.44) and 2015/16 (APR: 1.68; 95% CI: 1.36, 2.09) surveys respectively. Also, after adjusting for other factors, adolescent girls and young women with two or more live births (multipara) had 2.10, 2.42, 1.87 significantly higher prevalence of unmet need for modern contraception compared to those with no livebirth (nulliparous) in 2004/05 (APR: 2.10; 95% CI: 1.60, 2.75), 2010 (APR: 2.42; 95% CI: 1.76, 3.33) and 2015/16 (APR: 1.87; 95% CI: 1.43, 2.44) surveys respectively.

Furthermore, adolescent girls and young women who were sexually active during the past 4 weeks prior the survey had 59%, 68% and 55% significantly higher prevalence of unmet need for modern contraception compared to their counterparts in 2004/05 (APR: 1.59; 95% CI: 1.33, 1.91), 2010 (APR: 1.68; 95% CI: 1.36, 2.07) and 2015/16 (APR: 1.55; 95% CI: 1.28, 1.87) surveys respectively (Table **4**).

**Table 4:**
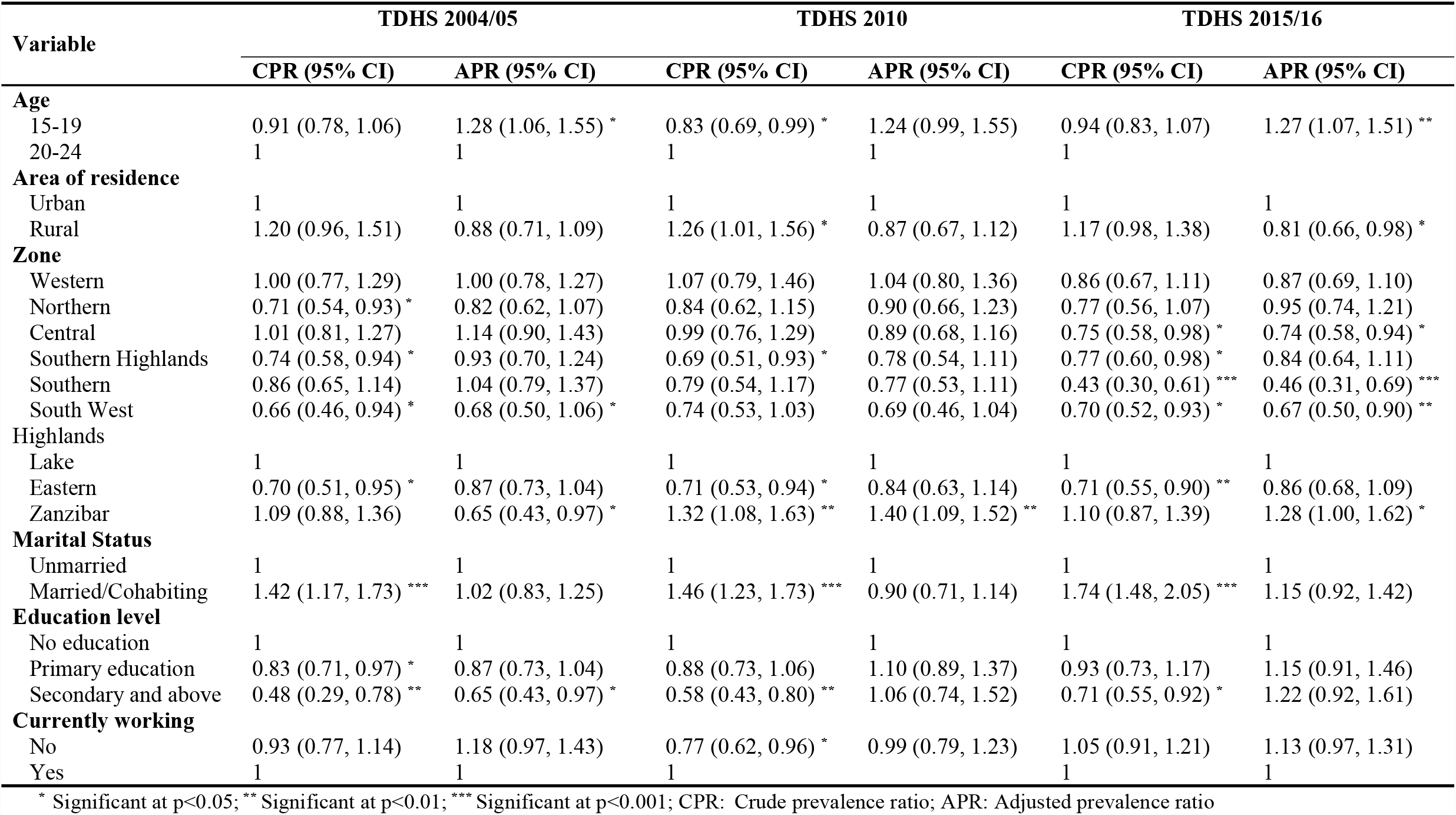
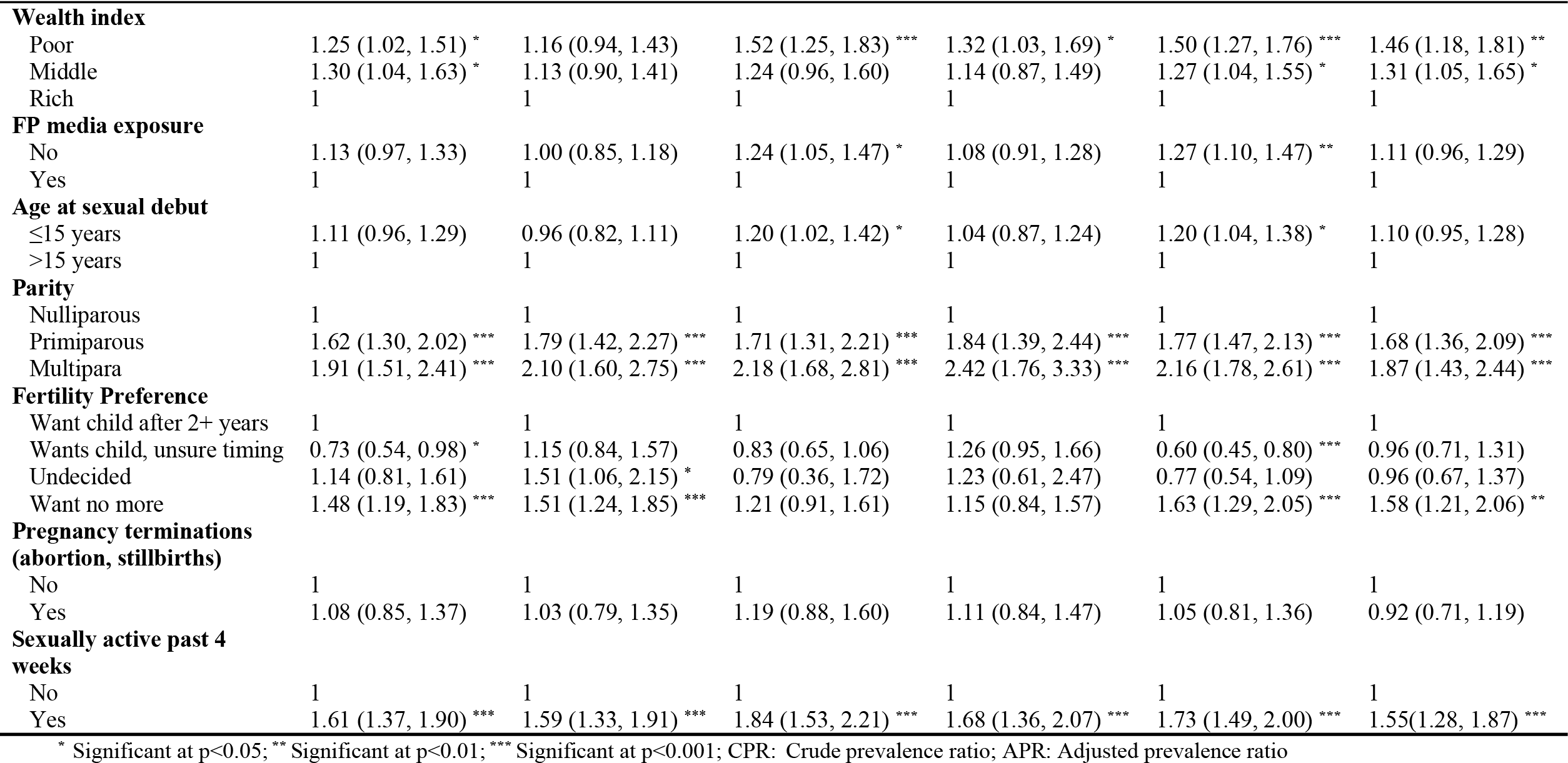
Crude & Adjusted analysis of Factors associated with unmet need for modern contraception among sexually active adolescent girls and young women in 2004/05, 2010 and 2015/16 (N=7,525)

## Discussion

The aim of this study was to determine the trends and determinants of unmet need for modern contraception among sexually active AGYW in Tanzania (2004-2016). We have shown that the unmet need for modern contraception declined by 4.3% over the 10-year period (31.8% in 2004/05 to 27.5% in 2015/16, respectively). Higher proportion of unmet need for modern contraception was observed among married compared to unmarried adolescent girls and young women. Numerous factors were associated with a higher prevalence of unmet need for modern contraception. These included younger age (15-19 years), poor wealth tertile, having at least one live birth, fertility preference of wanting no more children, being sexually active during the past 4 weeks prior the survey.

The decline in the trend of unmet need for modern contraception over the past 10-year period could mean that those AGYW who need modern contraception are able to access and use it. Similar findings have been reported from a study conducted in Ethiopia, as well as two DHS comparative studies (6,23,24). The observed decline in this setting could partly be explained by the increased national efforts to reposition family planning programmes. Additionally, a number of communities and schools based initiatives performed by different stakeholders through participatory sexual and reproductive health education, strengthened youth-friendly sexual and reproductive health services could also be a possible reason for the observed decline in unmet need for modern contraception (25–30).

The decline we present in the unmet need could also mean that there are less AGYW who need modern contraception, perhaps insinuating delays in sexual activity amongst this group. However, our further analysis demonstrated that younger age (15-19 years) was associated with a higher prevalence of unmet need for modern contraception, thereby suggesting there has been no delay in the onset of sexual activity. Evidence from previous studies in Tanzania report that more than 40% of adolescents are sexually active (31). Early age at sexual debut has been reported as an important predictor for unintended pregnancies which reflects unmet need for contraception (32). It has been widely reported that access to FP services for sexually active adolescents is restricted due to them being denied services, and also due to prejudices that health workers hold, including their preconceived ideas of what services adolescent girls should receive instead of providing the services actually needed (33,34). This compromises adolescents’ health seeking behavior thereby causing poor modern contraceptives uptake due to misinformation and fear of being called promiscuous as a result increased fertility rates and increased rates of unintended pregnancies in this age group.

We also found that AGYW from poor wealth tertile had higher unmet need for modern contraception. Previous studies have reported that women from poorer wealth quintiles are more likely to be less educated or out of school, unempowered, uninformed and thus less likely to access and utilize modern contraception (8,35–37). This also could be the reasons for the observed high unmet need for modern contraception among AGYW from poor wealth tertile in our study. This emphasizes that empowering and improving young women’ autonomy may help to reduce the rates of unmet need for modern contraception in this high-risk group.

Additionally, we also found that having at least one live birth is associated with higher unmet need for modern contraception. Evidence suggests that, postpartum contraception aids in facilitating optimal inter-pregnancy intervals therefore reducing the adverse maternal and child outcomes(38,39). Our finding suggests a need to scale up quality postpartum contraceptive services among married and unmarried AGYW especially among first time mothers.

This study has various strengths and limitations that should be considered when interpreting our findings. Our study benefited from a nationally representative large dataset which ensured adequate power and provided an opportunity for generalization of our findings. The application of Poisson regression model accounting for complex survey nature of the data enabled us to overcome the downfalls of classical logistic regression model when estimating common outcomes (>10%). However, the information obtained in this study was self-reported and subject to reporting bias and social desirability bias especially regarding the timing of most recent sexual intercourse especially among unmarried AGYW. Also, our study was cross-sectional in nature, we were therefore not able to infer causal-effect relationship because both exposures and unmet need for modern contraception (outcome) were measured at one point in time.

## Conclusion

Our study highlighted a steady declining trend of unmet need for modern contraception amongst AGYW in Tanzania from 2004 to 2016. Factors such as age, wealth index, parity, fertility preference and recent sexual activity have influenced unmet need over time. It is important to consider designing measurable targeted interventions in consideration of adolescent girls’ and young women’s needs and diversities. In particular, adolescents, AGYW from a poor wealth tertile, those with at least one live birth need to be empowered, and educated on their reproductive health rights and needs. This will further the uptake of modern contraceptive use for those in need and reduce the rates of unintended pregnancies, lower the adolescent fertility rate as a result lower unmet need for modern contraception. Eventually, helping Tanzania to accelerate progress in reaching national targets as well as SDGs of ensuring universal access to AYFSRHS including family planning, by 2030.

## Data Availability

Data are available from https://dhsprogram.com/Data/. DHS data may be accessed through registration of a research project on the DHS website and requesting access to data. This is the same manner the authors obtained access to the data. The authors did not have any special access privileges to DHS data

## Acknowledgement

We acknowledge the academic staff of Institute of Public Health-Kilimanjaro Christian Medical University College (KCMUCo), Epidemiology and Biostatistics KCMUCo class of 2020, Prof. Sia Msuya, Dr. Innocent Mboya for their tireless contribution during the course of this study.

## Abbreviations

AGYW: Adolescent girls and young women
TDHS: Tanzania Demographic and Health Survey

